# Stromal Tumor Infiltrating Lymphocytes (sTILs) as a putative prognostic marker to identify a responsive subset of TNBC in an Indian Breast Cancer Cohort

**DOI:** 10.1101/2020.08.19.20177865

**Authors:** Pooja M. Vaid, Anirudha K. Puntambekar, Rituja A. Banale, Ruhi R. Reddy, Rohini R. Unde, Namrata P. Namewar, Devaki A. Kelkar, LS Shashidhara, Chaitanyanand B Koppiker, Madhura Kulkarni

## Abstract

**Objectives:** Prognostic significance of stromal tumor infiltrating lymphocytes (sTILs) is evaluated to identify a responsive subset of TNBC in an Indian cohort of breast cancer patients.

**Methods:** A retrospective cohort of breast cancer patients from a single onco-surgeon breast cancer clinic treated with uniform treatment strategy across is evaluated for sTILs. FFPE tissue of primary tumor of invasive breast carcinoma are collected with ethical approvals. Tumor sections blinded for subtypes are stained with H&E and scored for sTILs by a pathologist following Immuno-Oncology TILs working group’s scoring guidelines.

**Results:** Analysis of 144 primary breast tumors for sTILs scores re-enforces significantly higher infiltration in TNBC tumors than HER2+ and ER+ tumors. Higher sTILs scores co-relate with gradually incremental pathological response to therapy specifically in TNBC subset and with better disease-free survival outcomes. Within TNBC, older and post-menopausal patients harbor higher scores of sTILs.

**Conclusion:** Despite a small cohort of breast cancer patients, TNBC subtype reflected significantly higher scores of sTILs with better response to therapy and disease-free outcomes as compared to other breast cancer subtypes. A larger number of breast cancer patients from an Indian cohort will strengthen the findings to establish sTILs as a marker to identify a responsive subset of TNBC.

**Key points:** *Key point 1:* This is a first attempt to understand the significance of stromal tumor-infiltrating lymphocytes in a breast cancer cohort from India, where higher recurrence and mortality rates are observed.

*Key point 2:* TNBC tumors show higher sTILs infiltration compared to non-TNBC patients. Higher sTILs scores within TNBC co-relates with better therapy response disease-free survival.

*Key point 3:* Higher sTILs scores in TNBC tumors in an Indian cohort showed a novel and unique association with old age and post-menopausal patients.

## INTRODUCTION

Breast Cancer is the leading cause for cancer related deaths in India, with close to 50% mortality rate^1^. Though there are effective treatments available for Breast Cancer, most targeted therapies are available for subtypes that express the molecular receptors ER, PR and/or HER2. A subtype that lacks expression of these receptors; triple negative breast cancer (TNBC) cannot avail targeted therapy^2^. With lack of targeted treatment, TNBC is an aggressive subset of breast cancer with higher rates of recurrence and lower overall survival^3,4^.

Worldwide, the prevalence of TNBC is 10–12%^5,6^, while in India, the prevalence of TNBC is reported to be significantly higher and up to 20–30%^7–9^. Moreover, the proportion of TNBC that presents with aggressive clinicopathological features such as younger and pre-menopausal age at incidence, high grade is also higher in Indian population^7^. With no targetable treatment and higher proportions of aggressive disease at incidence, TNBC poses a clinical challenge in India.

As of now, prognostication of TNBC is guided by clinicopathologic features such as tumour size, proliferative index, lymph node involvement as well as age and co-existing conditions. Standard treatment for TNBC includes neoadjuvant chemotherapy (NACT), followed by surgery^10–12^. TNBC is reported to show better response to NACT than ER+ve subtype i.e. 22% to 56% depending on the treatment regime used^3,13,14^. Pathological complete response in TNBC subtype is associated with better disease free survival^13,14^. Cases with residual disease have been shown to have a significantly higher chance of recurrence within 1^st^ three years of treatment and reduced overall survival^14–16^.

Recent studies have revealed tumor infiltrating lymphocytes (TILs) to be a promising prognostic marker to predict response to therapy, specifically in TNBC^17–19^. Tumor infiltrating lymphocytes are cytotoxic lymphocytes infiltrating into tumor and stromal regions as a host immune response to target cancer cells^20,21^. Higher proportion of infiltrating lymphocytes, especially in the stroma co-operate with the exogenous therapy enhancing the anti-tumor effects of the therapy^22,23^. Meta-analysis of 3770 patients with higher TILs scores predicted complete response to NACT in 50% of the TNBC patients and high TILs scores associated with better long-term survival over 3 years, emphasizing prognostic significance of TILs in TNBC^24,25^.

With high proportion of TNBC in India, understanding TILs distribution and its association with response to therapy and disease outcome may help predict a subset that is responsive and has better chance at disease free survival. If similar associations are found as seen in meta-analysis of western cohorts, TILs evaluation may provide a promise to predict a responsive subset of TNBC in India. In addition, TILs are easy to assess on histopathology of tumor tissue biopsies thus can be cost-effective and easily employable prognostic tool especially for low-resource countries like India.

Here we evaluate TILs with respect to clinicopathological features and outcomes of breast cancer patient cohort from a single surgeon and oncologist breast cancer unit in India. All the patients in the cohort have received similar treatment regimens and follow-up, giving high confidence in uniformity regarding treatment strategies. Primary, pre-treatment tumor tissue of 144 breast cancer patients are scored for stromal TILs according to the guidelines provided by the International TILs working group^23^. Association of TILs scores with clinicopathological features such as age at diagnosis, menopausal status, grade, tumor size, lymph node involvement and disease outcomes are evaluated. For patients that received NACT (n = 35), response to therapy and its association with TILs scores is also evaluated.

Despite the size of the cohort, a significant association of high sTILs scores with TNBC subtype is observed. Higher sTILs scores co-relate with better response to therapy and better disease outcome specifically in TNBC compared to non-TNBC. sTILs analysis within TNBC reflected distinct distribution between young, pre-menopausal patients as compared to old and post-menopausal patients. Old, post-menopausal patients presented with higher sTILs scores. Detailed co-relation with various clinical parameters of TNBC in comparison to non-TNBC is reported here.

## MATERIALS AND METHODS

### Patient tissue samples and meta-data

Formalin-fixed paraffin-embedded (FFPE) tissue samples and associated deidentified patient meta data are received from the clinic, following appropriate patient consent and ethical approval (dated 21st July 2018 #IECHR/VB/2018/016). Patients who were diagnosed at or underwent/are undergoing treatment at the clinic from 2012 up to 15^th^ March 2019 are included in the study. Patient data, including diagnostic clinico-pathological data, type of therapy and post-treatment follow-up data up to last follow-up /recurrence date/death is compiled and digitized. All FFPE tissue blocks used are of primary (pre-treatment) tissue. Of the 144 FFPE tissue samples used in this study, 106 are prepared from core biopsy tissue. 61 samples are of the naive tumor tissue block prepared from surgically excised tissue. All surgery tissue used is from patients who did not receive neo-adjuvant therapy.

Molecular subtypes are inferred based on patient immunohistochemistry reports. Samples are divided into ER+/HER-, HER+ and TNBC based on ER/PR expression and HER2 scores. ER+/HER- samples are with more than 1% ER expression and HER2 IHC score of 0, 1 or 2 and FISH-negative. Samples with IHC HER2 grade 3 or grade 2 FISH positive are taken as HER2+. Triple-negative samples are the ones with less than 1% ER and PR expression and HER2 IHC grade 1 or 2 and FISH-negative. Pre-treatment primary tumor tissue selection for these three molecular subtypes is described in the flowchart in Figure 1.

**Figure 1:**
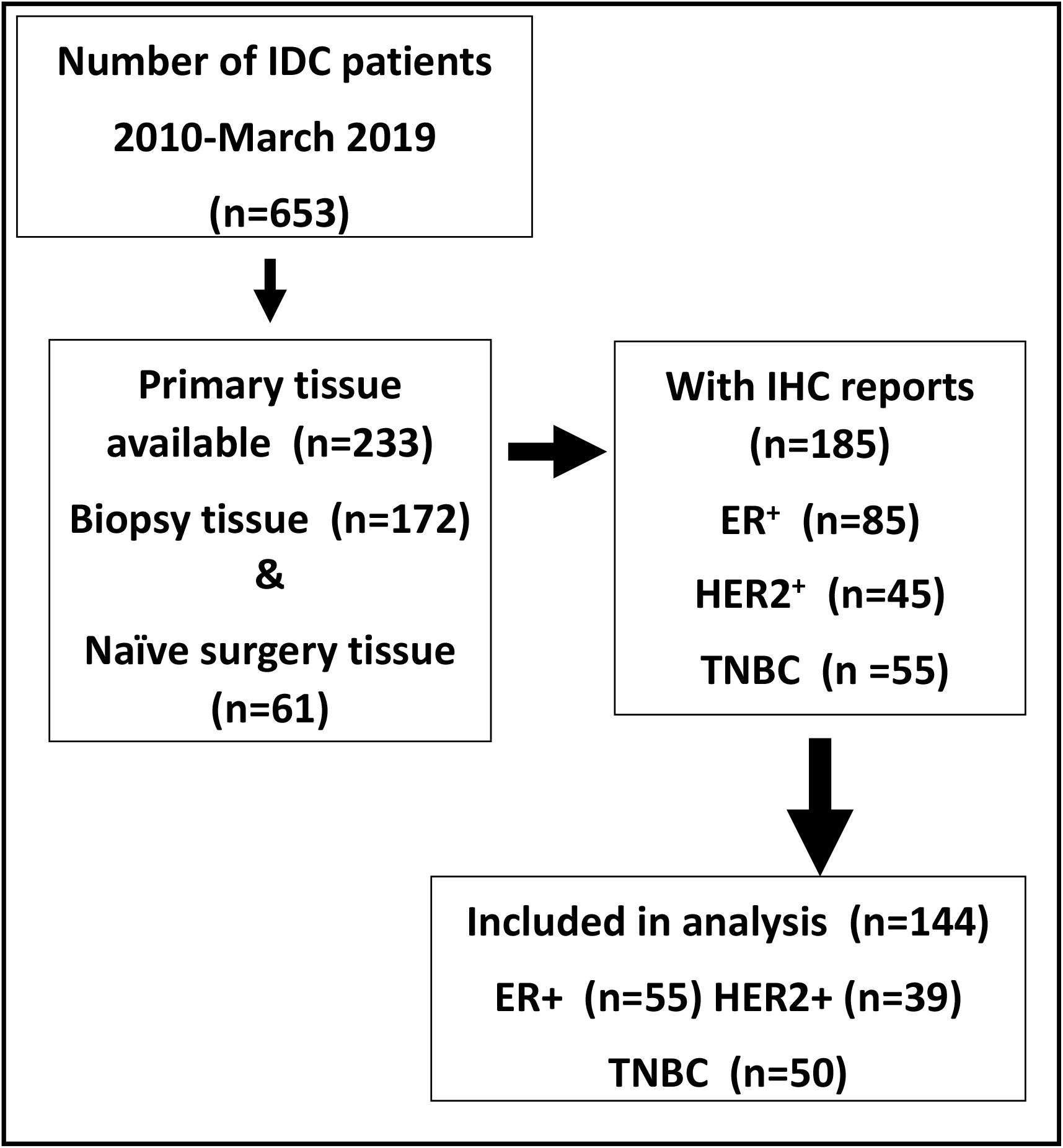
Flow chart to explain selection of primary breast cancer tissue. Primary tumor tissue selection from retrospectively (2010-March 2019) collected PCCM biobank. Out of 653 patients breast tissues deposited with patient consent at PCCM biobank, 233 were of primary tumor (172 biopsy tissue and 61 naïve surgery tissue). Out of these, molecular subtypes were assigned for 185 for whom IHC reports were available. Out of 185, 144 tissue samples with good tissue integrity were sectioned, stained with H&E and then scored for stromal tumor infiltrating lymphocytes following International TILs working group guidelines.

For NACT, TNBC patients (n = 14) were treated with Taxanes with or without an Anthracycline/Cyclophosphamide (AC) or 5-Fluoro-Uracil/AC regimen. ER+ patients (n = 15) were treated with anti-estrogens such as Letrozole or Tamoxifen. For HER2 positive patients (n = 6), AC followed by a Taxane with trastuzumab was administered.

For ACT, most TNBC cases received AC + Taxane as adjuvant therapy, some cases received Fluorouracil in addition. For non-TNBC cases, HER2+ cases were mostly treated with Trastuzumab with or without a Taxane, or FAC and Taxane, some hormone positive HER2 cases received hormonal therapy. Hormone positive cases were mostly treated with hormone therapy or with an AC + Taxane regimen.

Response to NACT is computed for 33 out of 35 patients who underwent NACT. Post-NACT, response to treatment is calculated by comparing cTcN values with ypTyPN. In case of absence of residual tumor and absence of lymph node metastasis during pathological examination of surgically removed tissue i.e. ypT0N0 status, response is considered as pathologically complete response (pCR). The presence of residual tumor and/or lymph node metastasis in surgically removed tissue is referred as Residual disease (RD). Response in patients with residual disease is further subdivided into three categories – partial, stable or progressive disease when compared to pre-treatment tumor size and lymph node positivity (stage). Partial response cases show downstaging, stable disease has no change in stage and progressive disease shows an increase in tumor size and/or lymph node positivity.

#### Histopathology of FFPE tissue blocks

FFPE blocks once selected are processed for histopathology. Tumor sections of 4–5 µm are obtained using Leica Microtome RM2255. Tissue slides are deparaffinized. Each slide is cleaned and stained with a drop of undiluted Hematoxylin solution (Delafield, 38803) in a humidifying chamber for 15 mins followed by 1% eosin (Qualigen Q39312). The slides are then gradually dehydrated in ethanol solutions followed by Xylene. Slides are mounted in DPX (Q18404).

#### Imaging of histopathology slides

All slides are imaged by OptraScan using OS-15 bright field digital scanner at 400X magnification. Images are viewed using the ‘Image viewer’ software provided by Optra. These are then converted to TIFF format and processed using Image Viewer Version 2.0.4 by OptraScan for scale bars.

#### Stromal Tumor Infiltrating Lymphocytes (sTILs) Scoring

TILs percent distribution within stroma surrounding the tumor tissue is assessed from H&E stained histopathology section of pre-treatment primary tumor tissue. The scoring is done by a single pathologist (AP) according to the recommendations of The TILs Working group^23^. The pathologist was blinded to the clinical data as well as molecular subtypes of the tissue. For our study, only stromal TILs are scored and used for the analysis.

#### Statistical Analysis

All statistical analysis is done using GraphPad Prism v.5. Demographic table was prepared using IBM SPSS Statistics v. 21.0.0.0. Distribution of clinicopathological characteristics within the cohort and breast cancer subtypes is tested using 2×2 (2×4 in case of tumor size) Chi square contingency test. Mean age across subtypes is compared using unpaired two-tailed Student’s t-test. Column statistics for sTILs are computed on GraphPad Prism v.5 to calculate mean and SD/SE across each sub-category of clinical characteristics. Significant difference in distribution between TNBC and on-TNBC sTILs scores across clinicopathological characteristics is tested with one-way ANOVA followed by Tukey’s Multiple Comparison Test. Statistical analysis is done using GraphPad Prism v.5 and graphs are plotted.

Disease outcomes are computed as follows: disease free survival (DFS) is calculated as time in months from date of surgery till date of recurrence or last follow-up date. Overall survival (OS) is calculated as time in months from the diagnosis date (biopsy date) till last follow-up date or date of death due to disease. Kaplan-Meier survival plots for DFS and OS for up to 3-years follow-up time are plotted and survival probabilities are computed by Log-rank, Breslow and Tarone-Ware cores towards 3-year DFS and OS.

Distribution of response to NACT and breast cancer subtypes is tested using 2×4 Chi square contingency test. Box plot for sTILs distribution across response to NACT i.e. pCR and RD is plotted on GraphPad Prism v.5. Mean sTILs and SE for different response to NACT i.e. complete, partial, stable or progressive is computed and plotted by using GraphPad Prism v.5. Mean sTILs for different response to NACT across breast cancer subtypes is calculated and plotted using GraphPad Prism v.5.

## RESULTS

### Breast Cancer Cohort Characteristics

Pre-treatment primary tumor tissues were identified for 144 patients from the biobank as described in Figure 1. Demographic and clinical characteristics of the entire cohort and according to their molecular subtypes (TNBC and non-TNBC) are presented in Table 1. Average age of the cohort of 144 breast cancer patients is 53 ± 12 years with a range from 28–86 years. TNBC patients (n = 50) with mean age of 50, showed equal distribution within young (< 50 years) and late (> 50 years) age. ER+ve (n = 55) and HER2+ve (n = 39) patients presented higher mean age of 56 and 54 years with higher proportion (69% and 64%; respectively) in late age category. 62% TNBC patients presented at postmenopausal age, and menopausal status is not significantly different across the subtypes. ER+ and HER2+ patients show significantly higher proportion of low grade (89% and 62%; respectively) tumours, while TNBC shows close to equal distribution within low and high tumor grade (45% and 55%). Other than grade, tumor size (cT, pT), lymph node status (cN, pN), lymph-vascular invasion (LVI) do not show significantly different distribution across all three subtypes. Overall survival and disease free survival data is available for 131 patients, with average follow-up of two years ranging from 0 months to 14.3 years, and median follow-up of 1.7 years.

**Table 1:** Demographic and clinical features of breast cancer patient cohort. Number of patients with breast cancer were grouped into, ER+/PR+/PR-/HER2-, HER2+ and TNBC. The number of patients are listed according to the clinical variables reported at the time of diagnosis as per both the subtypes. Clinical parameters such as age at diagnosis, menopausal status, tumour grade and radiological tumor size, lymph node positivity and LVI are listed. For patients who did not receive NACT/NAHT, pT and pN retrieved from the surgery pathology report are noted. For patients who received NACT/NAHT, pathological response to the therapy is noted based on their ypTN status. Total number of patients with follow-up and time to follow-up and follow-up status are also noted. Any skewed distribution of clinical parameters in the cohort was tested using the 2*3 (4*3 in case of tumor size) χ2 contingency test with GraphPad Prism v.5. Red font indicates significant p-values *For comparing mean age differences among the subtypes, 1-way ANOVA was used. LVI- lympho-vascular invasion, Neo-adjuvant chemotherapy (NACT), pCR- pathological complete response.

|  |  | BC | ER+ | HER2+ | TNBC | p-values |
| --- | --- | --- | --- | --- | --- | --- |
| No. of patients |  | 144 | 55 | 39 | 50 |  |
| Age<br>(n=144) | (Mean ± S.D) | 53.4 ± 12.3 | 55.9 ± 12.4 | 54.2 ± 11.6 | 50.0 ± 12.2 | *0.0455 |
|  | Early (>50) | 56 | 17(30.9%) | 14(35.9%) | 25(50%) | 0.1214 |
| | Late (≤ 50) | 88 | 38(69.1%) | 25(64.1%) | 25(50%) | |
| Menopausal<br>status<br>(n=129) | pre | 35 | 9(20%) | 8(21.6%) | 18(38.3%) | 0.0958 |
|  | post | 94 | 36(80%) | 29(78.4%) | 29(61.7%) |  |
| Grade<br>(n=140) | Low (I/II) | 92 | 46(88.5%) | 24(61.5%) | 22(44.9%) | < 0.0001 |
|  | High (III) | 48 | 6(11.5%) | 15(38.5%) | 27(55.1%) |  |
| Tumor size<br>(cT)<br>(n=134) | T1 | 35 | 21(40.4%) | 7(19.4%) | 7(15.2%) | 0.054 |
|  | T2 | 93 | 28(53.9%) | 28(77.8%) | 37(80.4%) |  |
|  | T3 | 4 | 2(3.8%) | 0(0%) | 2(4.4%) |  |
|  | T4 | 2 | 1(1.9%) | 1(2.8%) | 0(0%) |  |
| LN status<br>(cN)<br>(n=129) | negative | 59 | 25(53.2%) | 14(38.9%) | 20(43.5%) | 0.4011 |
|  | positive | 70 | 22(46.8%) | 22(61.1%) | 26(56.5%) |  |
| pT (primary<br>tissue, no NACT)<br>(n=95) | T0 | 5 | 3(9.7%) | 0(0%) | 2(6.1%) | 0.3852 |
|  | T1 | 17 | 8(25.8%) | 4(12.9%) | 5(15.1%) |  |
|  | T2 | 68 | 18(58.1%) | 26(83.9%) | 24(72.7%) |  |
|  | T3 | 4 | 1(3.2%) | 1(3.2%) | 2(6.1%) |  |
|  | T4 | 1 | 1(3.2%) | 0(0%) | 0(0%) |  |
| pN (primary<br>tissue, no NACT)<br>(n=95) | negative | 65 | 23(74.2%) | 19(61.3%) | 23(69.7%) | 0.54 |
|  | positive | 30 | 8(25.8%) | 12(38.7%) | 10(30.3%) |  |
| LVI<br>(n=130) | negative | 95 | 39(76.5%) | 26(70.3%) | 30(71.4%) | 0.777 |
|  | positive | 35 | 12(23.5%) | 11(29.7%) | 12(28.6%) |  |
| NACT<br>(n=141) | No | 106 | 37(71.2%) | 33(84.6%) | 36(72%) |  |
|  | Yes | 35 | 15(28.8%) | 6(15.4%) | 14(28%) |  |
| PCR status after<br>NACT<br>(n=33) | Complete | 8 | 1(7.1%) | 1(20%) | 6(42.9%) | 0.0855 |
|  | No response | 25 | 13(92.9%) | 4(80%) | 8(57.1%) |  |
| Survival outcomes | No. followed-up | 131 | 46 | 36 | 49 |  |
|  | Time in Months<br>Mean(range) | 25(0-172) | 20(0-84) | 28(0-172) | 28(1-132) |  |
|  | Median months | 19 | 12 | 26 | 20 |  |
|  | # Recurred<br>(local, distant) | 11 | 3 | 2 | 6 |  |
|  | # Death due to<br>disease | 3 | 0 | 0 | 3 |  |

To assess tumor response to NACT, patients who received NACT prior to surgery are identified. Out of 144 patients, 35 (24%) had received neoadjuvant chemotherapy (NACT) according to their molecular subtypes as described in ‘Methods’ section. Of these 35, 8 patients show complete pathological response (pCR) as assessed by ypT0ypN0 status.

### Stromal TILs Scores Distribution Across Clinical Parameters

Stromal TILs (sTILs) distribution score is evaluated for each tissue sample from H&E stained histopathology slide. Table S1A depicts the distribution of sTILs percent distribution across clinical features of the cohort. Average sTILs scores show even distribution across age, menopausal status, and lymph node status. Tumor grade and tumor size (cT and pT) show significantly skewed sTILs distribution where higher grade and larger size tumors are associated with higher sTILs percentages^26–28^.

### Stromal TILs Scores and Co-relation with Molecular Subtypes

sTILs scores are compared between TNBC, ER+ve and HER2+ve tumor tissue. Representative images of each histopathology sections for each subtype are shown in Figure 2A. TNBC tumors harbor wider range of sTILs scores (5 – 90%) as compared to other subtypes (0–60% and 0–70% for ER+ve and HER2+ve tumors) as shown in the box plot (Figure 1B). Mean sTILs score is significantly higher in TNBC (35.02 ± 3.93, n = 50) compared to ER+ve and HER2+ve tumors (11.02 ± 1.49, n = 55 and 21.64 ± 3.17, n = 39). sTILs scores are binned into three categories: Low (< 10%), Moderate (10–40%) and High (> 40%) (Figure S1A) according to International TILs Working Group 2014 guidelines for TILs evaluation^23^. Binned scores also reflect significantly skewed distribution between TNBC and other two subtypes. Less than 5 and 10% of ER+ve and HER2+ve samples refelct high TILs scores while close to 40% of TNBC tumors harbor high TILs scores. (Figure S1B).

**Figure 2:**
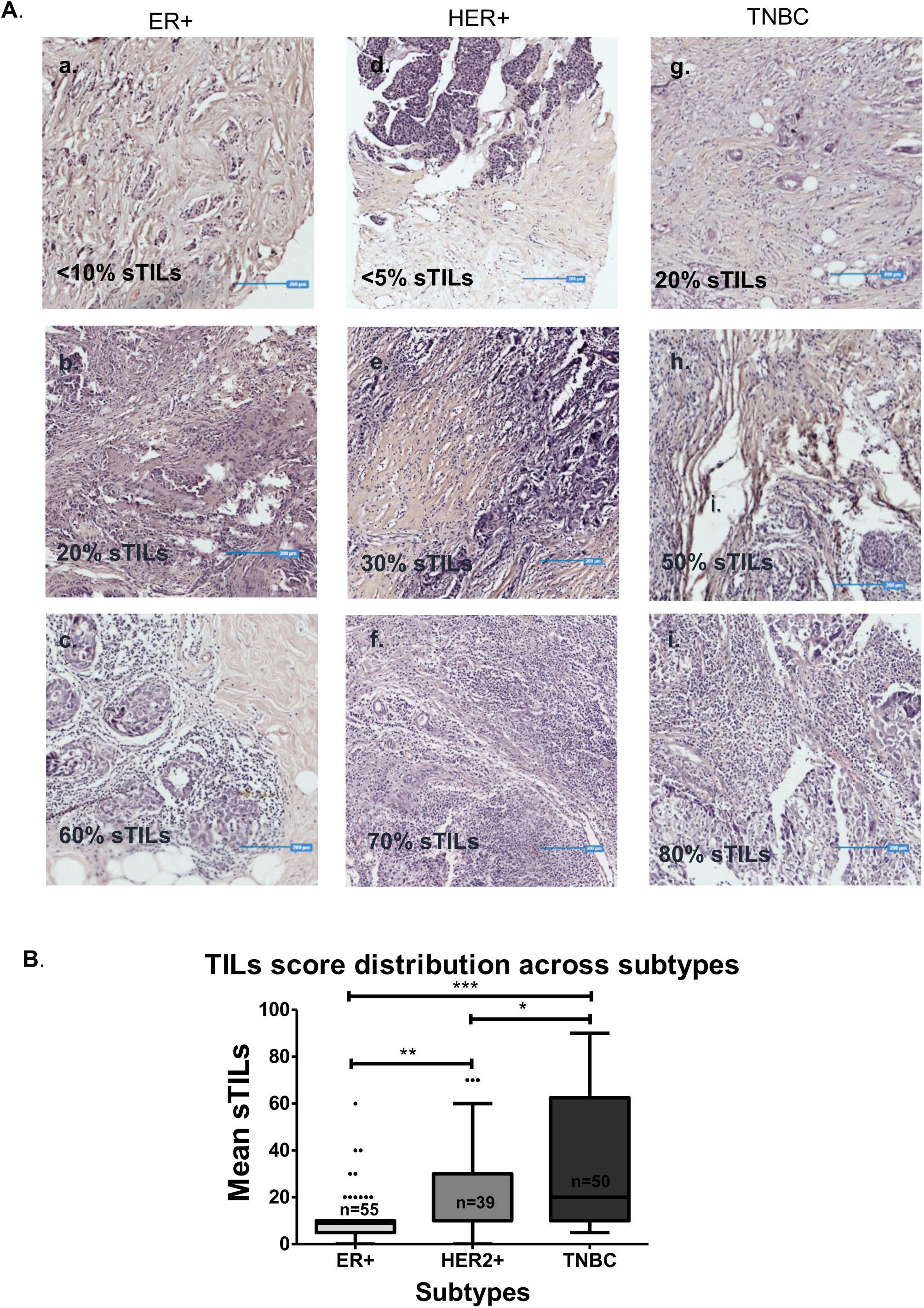
sTILs distribution across all three subtypes tumor tissue. **A**. Images depicting stromal TILs distribution. Representative images of sTILs scores for ER+(left panel), HER2+(middle panel) & TNBC (right panel). Percent sTILs score for each of the tissue is mentioned on the figure. Blue lines are the scale bars representing 200um. **B**. Box plot shows distribution of sTILs scores across ER+, HER2+ and TNBC tumor tissue. The horizontal line represents median. Error bars represent Tukey’s whiskers. The actual number of patient samples (n) are shown in the box plot. The distribution of the sTILs scores between tissue samples across each subtype was tested for statistical significance with Student’s t-test, using GraphPad Prism v.5. * represents p-value < 0.05, ** represents p-value < 0.01, and *** represents p-value < 0.0001.

sTILs scores are compared between the subtypes according to their clinical parameters (Table 2). Older age (>50) TNBC tumors reflect significantly higher proportion of sTILs scores compared to ER2+patients; 38.72 ± 5.83, n = 25 vs 10.74 ± 1.43, n = 38 respectively (Table 2 and Figure 2A). As expected, post-menopausal TNBC patients show significantly higher proportion of sTILs scores as compared to ER2+ve patients; 37.86 ± 5.39,n = 29 vs 10.08 ± 1.42, n = 36 respectively (Table 2 and Figure 2B). No significant difference was seen across mean sTILs between HER2+ve and TNBC patients. Overall distribution of sTILs through low and high grades across subtypes is significantly different with high grade TNBC tumors presenting with higher mean sTILs score; 42.56 ± 5.70 (n = 27), followed by high-grade HER2+ve patients; 31.00 ± 5.35 (n = 15) and ER+ve patients; 17.50 ± 3.59 (n = 6). (Figure 2C). Mean sTILs scores across clinical (cT) and pathological (pT) tumor size, lymph node (cN and pN) status showed higher scores for larger and lymph node negative tumors, specifically in TNBC as compared to non-TNBC subtypes (Figure 3D-G). Similarly, sTILs scores are significantly higher in LVI negative TNBC tumors as compared to ER+ve tumors (33.73 ± 5.27, n = 30 vs 11.05 ± 1.79, n = 39) as represented in box graph (Figure 3H).

**Table 2:** Mean sTILs scores with respect to clinicopathological features of breast cancer subtypes. Mean ± S.E sTILs scores across clinicopathological parameters including age at diagnosis, menopausal status of patients, tumor grade, radiological and pathological tumor size and lymph node status and LVI according to the molecular subtypes are presented. The statistical analysis was done in GraphPad Prism v.5. One-way ANOVA was performed to compute significant difference in mean sTILs scores between breast cancer subtypes and for comparing mean sTILs scores across each category and subtypes. Red font indicates significant p-values. *LVI- lympho-vascular invasion

**Figure 3.**
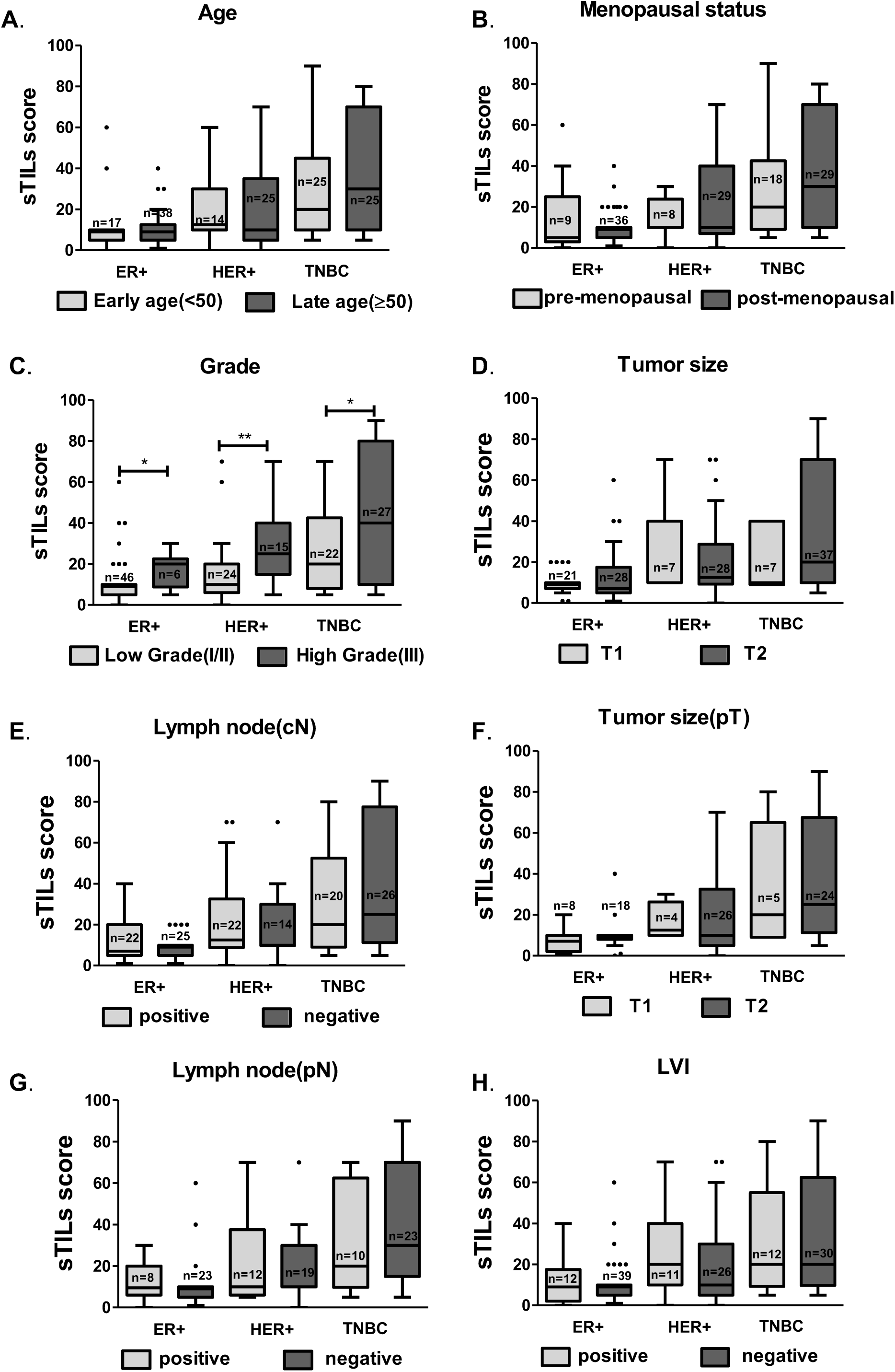
sTILs distribution across clinico-pathological parameters with respect to subtypes. Box plots depicting mean sTILs scores for ER+, HER2+ and TNBC tumor tissue across **A**. early & late age, **B**. pre & post-menopausal status, **C**. low (I/II)and high (III) grade tumors, **D**. clinical tumor size; T1 and T2, **E**. clinical lymph-node status – positive and negative, **F**. pathological tumor size; T1 and T2, **G**. pathological lymph-node status – positive and negative, **H**. LVI status – positive & negative. The number of tissue samples (n) are shown on top of each bar. Error bars represent Tukey’s whiskers. Unpaired t-test was performed to test for significance across each parameter between individual subtypes. Only grade shows significant differences across all three subtypes, which is represented here. p-value < 0.05 is represented with ‘*’, < 0.01 with ‘**’ and, < 0.0001 with ‘***’. ns = non-significant. GraphPad Prism v.5 was used for the graphs and ANOVA test.

### sTILs Scores and response to NACT

Out of 144 breast cancer patients, 35 patients received NACT. For 33 patients, pre and post-surgery records were available for cTcN and ypTypN data. ypT0ypN0 was taken as pathologically complete response (pCR). Out of 33, 8 patients show pCR post-NACT. Patients with complete response show wider range of sTILs scores compared to the rest who had residual disease (RD) post-NACT (n = 25) (Figure 4A). For the patients with residual disease, their response to NACT is further segregated into ‘partial response’ when the tumor is down staged by therapy, ‘stable disease’ when no change in the stage occurred post-NACT, and progression in the stage after NACT is taken as ‘progressive disease’ (Table S2). When sTILs scores are compared for each of these categories, inverse pattern is observed with treatment response. Patients with complete pathological response had highest mean sTILs scores of 30 ± 33 followed by partial response (22.90 ± 19.96) and stable disease (14.36 ± 15.60), and progressive disease with the least sTILs score of 5% (Figure 4B). sTILs scores for TNBC patients show similar gradual reduction for patients showing complete response (36.67 ± 36.01), followed by partial response (25.80 ± 26.23), and goes lower for patients with stable disease (10.00 ± 8.66) (Figure 4F).

**Figure 4:**
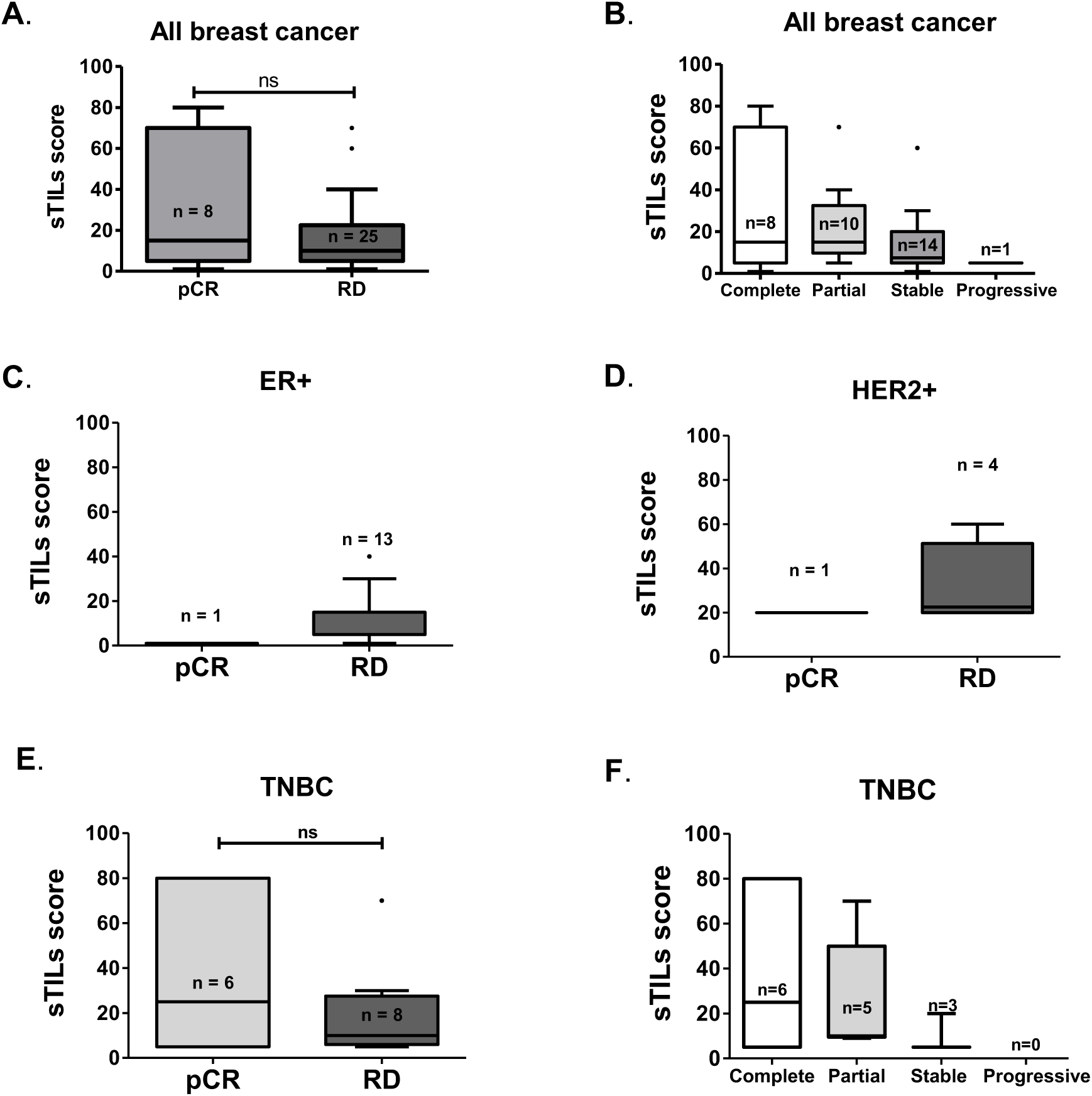
Mean sTILs scores and its association with response to NACT. **A**. sTILs score distribution is plotted for patients who showed complete pathological response (pCR) or had residual disease (RD) post-NACT. Box plots with horizontal lines show median sTILs score and The horizontal line represents, and error bars represent Tukey’s whiskers. Mean sTILs across both groups are not significantly different as tested by two-tailed unpaired t-test. **B**. Box plot representing mean sTILS score for all the patients that received NACT and showed either complete orpartial pathological response or had a stable or progressive disease at the time of surgery. Error bars represent Tukey’s whiskers. **C**. Mean sTILs scores for ER+ patients who had pCR or residual disease (RD). No statistics could be computed. Error bars represent Tukey’s whiskers. **D**. Mean sTILs scores for HER+ patients who had pCR or residual disease (RD). No statistics could be computed. Error bars represent Tukey’s whiskers. **E**. Mean sTILs scores for TNBC patients who had pCR or residual disease (RD). Unpaired t-test was run for sTILs score across pCR and RD. Error bars represent Tukey’s whiskers. **F**. Box plot representing mean sTILS score only for TNBC patients that received NACT and showed either complete or partial pathological response or had a stable or progressive disease at the time of surgery. Error bars represent Tukey’s whiskers. *pCR-pathological complete response, RD-Residual disease

For the patients who received NACT, sTILs scores and response to NACT are further co-related across subtypes. TNBC patients who show complete pathological response had higher mean sTILs score (36.67%) compared to the rest (20%) (Fig 4E). Irrespective to the response to treatment, ER+ve and HER2+ve patients had lower mean sTILs scores (Figure 4C and 4D).

### sTILs Scores and Disease Outcome

sTILs scores binned into three categories – low (< 10%), moderate (10–40%) and high (>40%) are analyzed for association with the disease outcome using Kaplan Meier curves. The cohort shows no specific association of binned TILs scores with disease free survival (Figure 5A). When subtypes are segregated differing outcomes for disease free status with respect to binned sTILs scores is reflected depending on the subtype. High sTILs scores in TNBC tumors are associated with longer relapse free life as compared to low or moderate TILs scores, with hazard ratio of 0.49 by Cox regression (Figure 5D). On the other hand, high sTILs shows significantly poor survival in HER2+ve patients. (Figure 5C), while ER+ve patients show similar trend as TNBC patients. For overall survival, the cohort presented with 3 deaths due to disease, and all of which harbored moderate sTILs scores (Figure 6).

**Figure 5:**
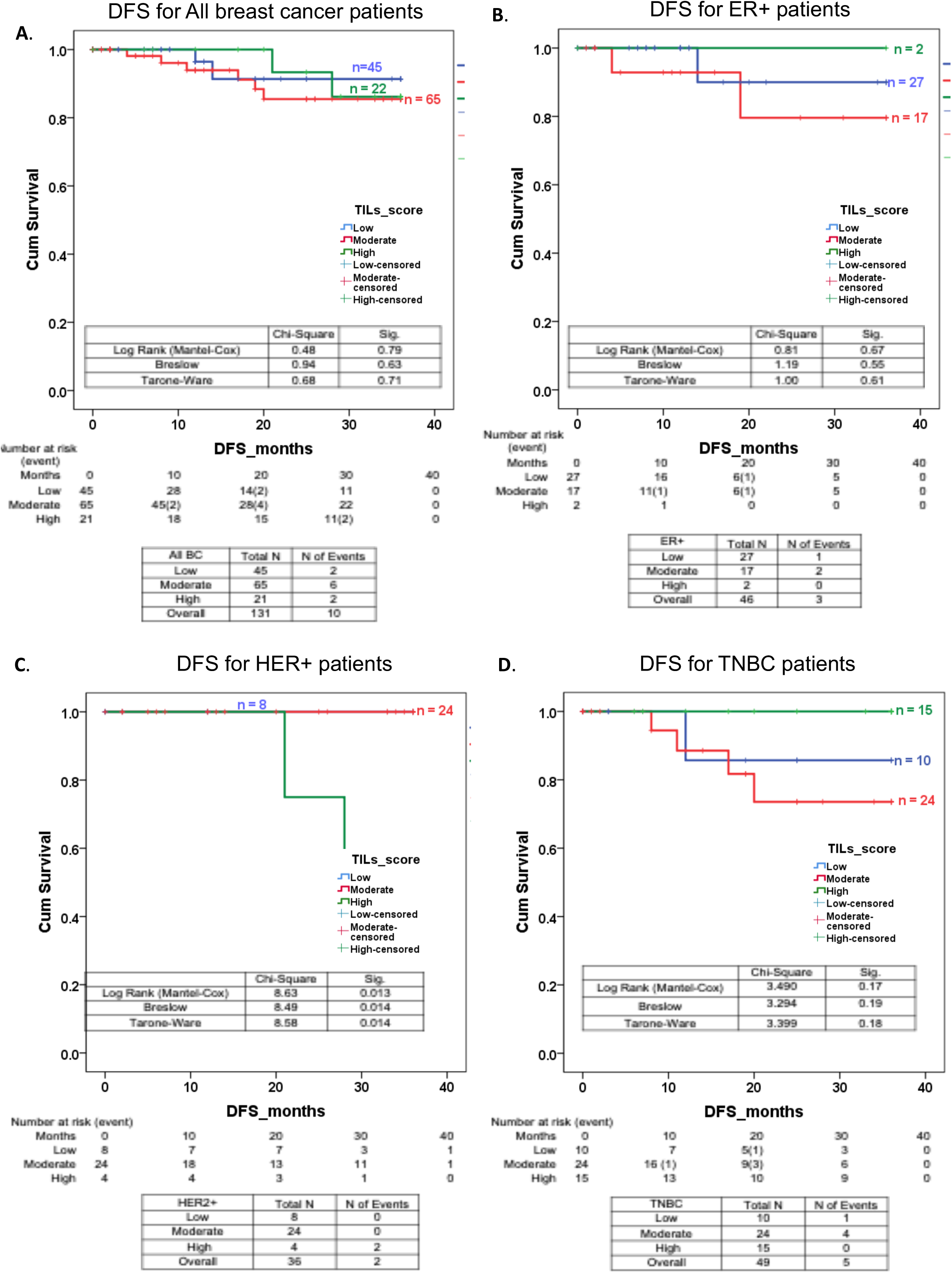
Three year disease-free survival with respect to binned sTILs scores. Disease free survival was calculated as number of months from the date of surgery till the recurrence diagnosis date or last follow-up date. The cut-off for DFS calculation was taken at 3- years and calculated for a total of 131 patients. Kaplan-Meier survival plots for disease-free survival (DFS) according to low, moderate & high sTILs score bins for A. the entire cohort, B. ER+ patients and C. HER2+ patients and D. TNBC patients. In the graph, X-axis represents time scale in months, Y-axis represents the survival probability. Blue-lines indicate patients with low sTILs score, red-line indicates patients with moderate score & green indicates high sTILs scores. Each drop shown as vertical line represents an event i.e. local or distant recurrence. Survival probability with respect to sTILs binned scores is tested using IBM SPSS Statistics v. 21.0.0.0.

**Figure 6:**
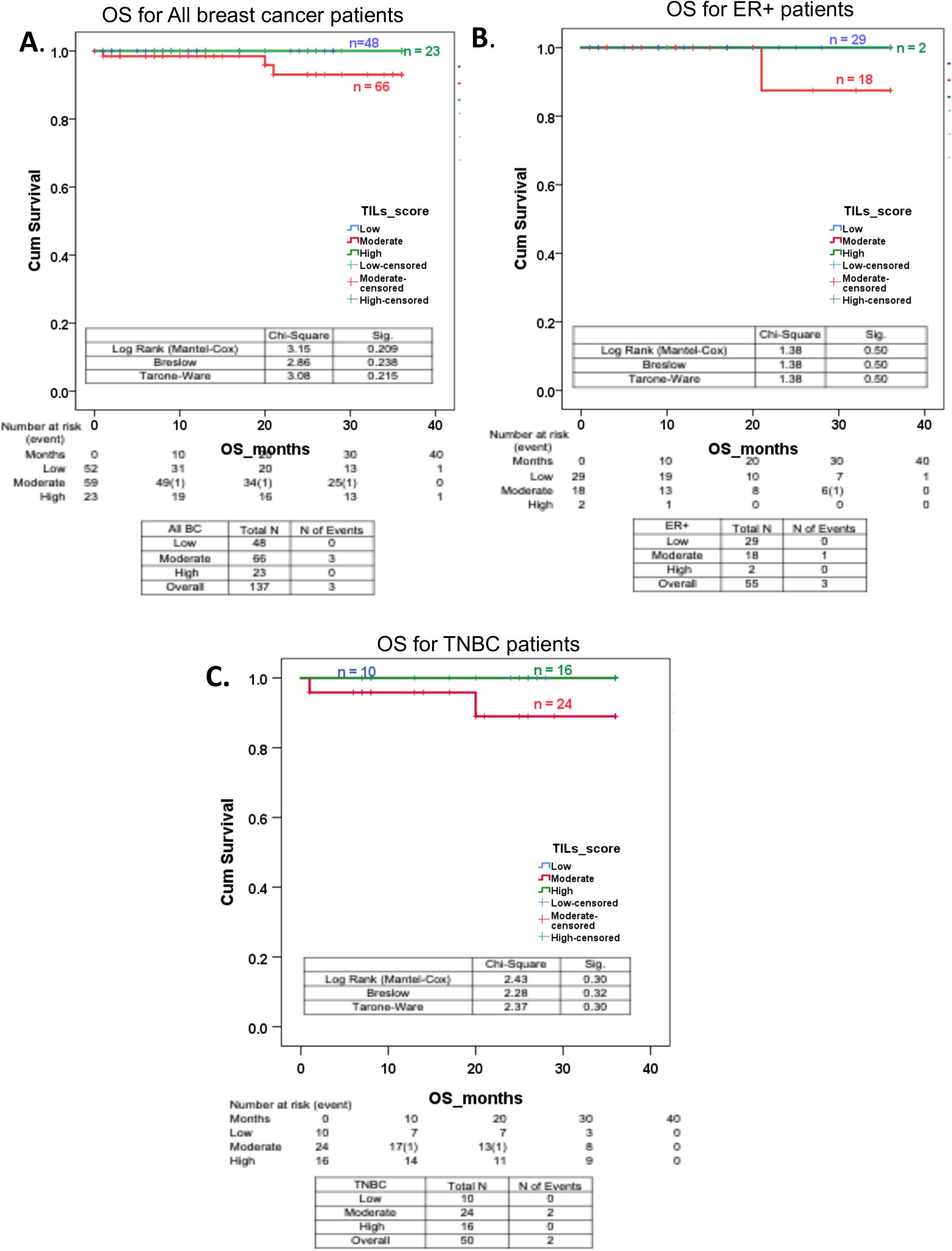
Three year overall survival with respect to binned sTILs scores. Overall survival was calculated as number of months from the date of biopsy till the last follow-up date. The cut-off for OS calculation was taken at 3- years and calculated for a total of 137 patients. Kaplan-meier survival plots showing overall survival (OS) of patients according to low, moderate & high sTILs score bins for A. the entire cohort and B. ER+ patients and C.TNBC patients. In the graph, X-axis represents time scale in months, Y-axis represents the survival probability. Blue-line indicates patients with low sTILs score, red-line indicates patients with moderate score & green indicates high sTILs scores. Each drop shown as vertical line represents an event i.e. death due to disease. Survival probability for each factor is tested by using IBM SPSS Statistics v. 21.0.0.0.

## DISCUSSION

This report is the first attempt to understand association of stromal TILs with clinicopathological features of breast cancer, their outcomes and response to therapy for an Indian cohort. The study is limited with a small cohort of 144 breast cancer patients, nevertheless the analysis confirms established association of sTILs with response to therapy and disease-free outcomes specifically in TNBC subtype. Further, this study brings forward significant co-relation of sTILs with clinicopathological features of TNBC subtype, perhaps specific to an Indian cohort.

The cohort of 144 breast cancer that contained 50 TNBC, 55 ER+ve and 39 HER2+ve patients reflected uniform distribution of clinicopathological parameters except for age and grade. TNBC presented with younger mean age as compared to non-TNBC and comprised of higher proportion (50%) of young age (< 50 years) and pre-menopausal patients, unique to an India Breast Cancer Cohort^7,9^. This is in contrast to the western cohorts where young age TNBCs present at 29–34%^29,30^. TNBC in our cohort also presented with higher proportion of lower grade tumors (45%) as compared to western cohorts (20.2%)^31^. For patients who received NACT (n = 33), 42.9% of TNBC patients had complete pathological response to the therapy as opposed to 7% and 20% of ER+ and HER2+ cohort. This is line with the current literature, where TNBCs indeed show better response to therapy^3,13,14^.

Stromal TILs scores in the cohort were uniformly distributed irrespective of the clinic-pathological parameters of the tumors, except for grade and tumor size. Higher grade and larger tumors showed significantly higher sTILs scores, as has been reported elsewhere^27,32^. Subtype wise comparison of sTILs scores co-related well with reported studies^23^ and meta-analysis^24^, where TNBC subtype presented with wider sTILs scores distribution with a greater number of tumors with high sTILs scores (35%) as compared to ER+ and HER2+ subtypes (11% and 22% respectively). Within TNBC, higher sTILs scores co-related with better disease-free outcomes. With the limited number of patients that received NACT, broader and higher sTILs scores associated with complete pathological response. The gradual reduction in response to therapy with gradual decrease in sTILs scores was confined to TNBC subtype, similar to that has been reported in western population^24^. The trends observed in our study are in line with the established association of sTILs distribution and TNBC outcomes, although not significant. Non-significant distribution patterns observed could be due to small cohort size or may be specific to Indian context, something that needs to be explored with larger cohort of breast cancer patients from India.

Apart from validating established associations of sTILs with disease outcome and response to therapy, our study uncovers unique associations of sTILs scores with clinicopathological features specifically in TNBC subtype. Significantly higher sTILs scores were observed for old age, post-menopausal TNBC patients as compared to young, pre-menopausal TNBC patients. This is in contrast to earlier reports, where a meta-analysis of nine studies showed significantly lower sTILs scores for older age^25^. While in a cohort of 897 TNBC patients, no significant difference in sTILs scores was noted between old and young age TNBC patients^33^. It is known in the field that young age TNBC patients, although with aggressive disease show better response to therapy and higher rates of pCR than older age TNBC patients^34,35^. Whether, higher sTILs scores in older age, post-menopausal TNBC patients of our cohort translates into better response to therapy and better disease outcome for them needs to be further analysed with a larger cohort. Perhaps such association could possibly turn out to be specific to TNBC in an Indian context.

Further analysis for sTILs distribution will provide validation to establish TILs as a predictive tool to segregate old age post-menopausal TNBC patients who may benefit from NACT and young age, pre-menopausal TNBC patients with aggressive disease who are most likely to be non-responders to the therapy.

## Data Availability

All the data is presented here in the manuscript. Further details can be provided upon request.

## Funding and Acknowledgement

MK would like to acknowledge DBT- Ramalingaswami ‘re-entry’ fellowship awarded by DBT-India. LSS would like to acknowledge DST-JC Bose Research Fellowship. Research grant to CTCR is supported by Bajaj Auto Ltd. PV would like to acknowledge Aditi Khatpe, Rutvi Shah and Dimple Adiwal for their help in standardization.

## Supplementary Legends

**Table S1:**
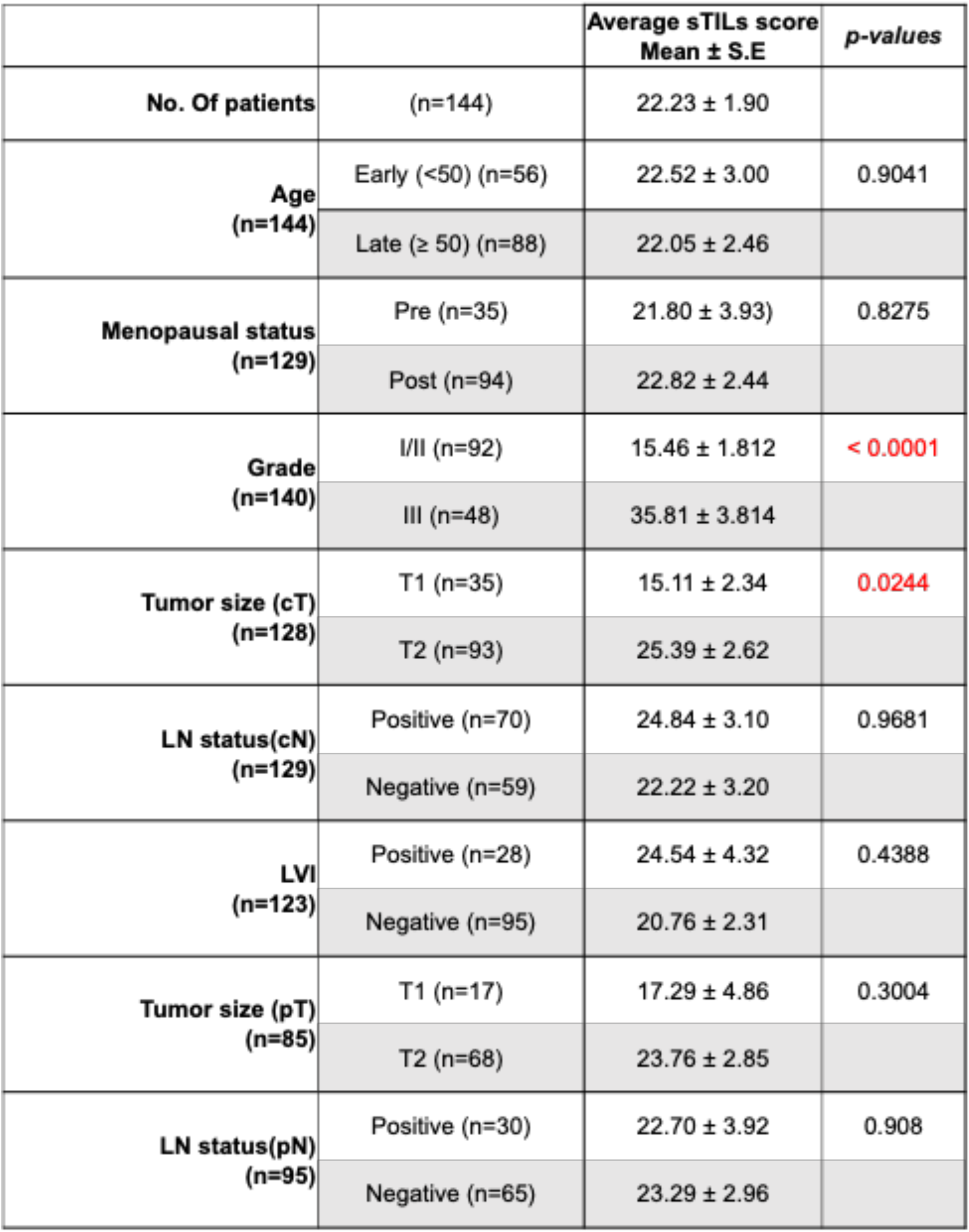
Mean sTILs scores with respect to clinical features of the cohort. Mean ± S.E of sTILs scores across clinicopathological parameters including age at diagnosis, menopausal status of patients, tumor grade, radiological tumor size and lymph node status and LVI. For comparing mean TILs score differences across each parameter two-tailed, unpaired t-test was performed on GraphPad Prism v.5. #Red font indicates significant p-values. * LVI- lympho-vascular invasion

**Figure S1:**
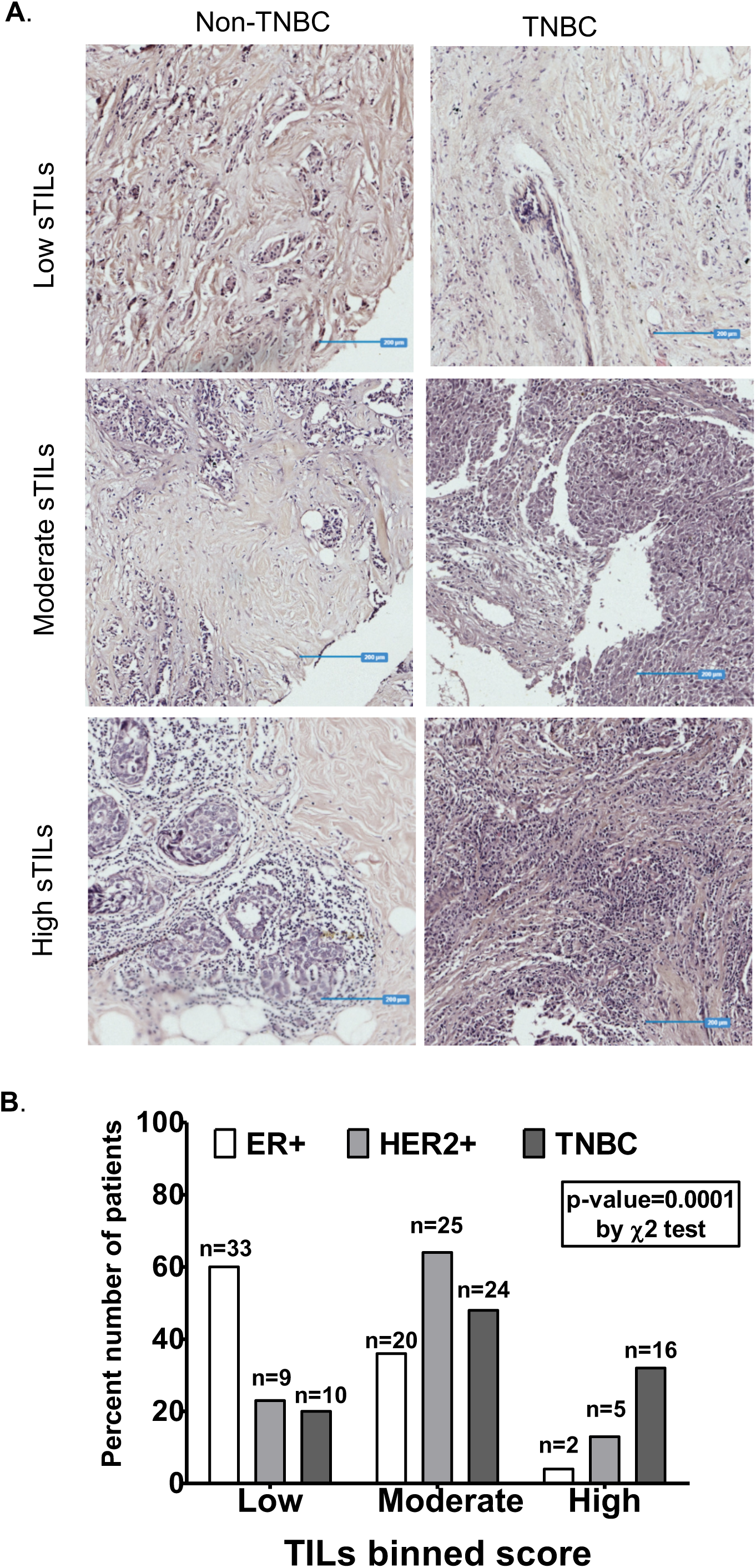
Binned sTILs distribution across all breast cancer subtypes tumor tissue. **A**. Representative images depicting stromal TILs distribution in tumor. Images of binned TILs scores are presented here for non-TNBC (left panel) & TNBC (right panel). Representative images of Low (< 10%), Moderate (10%-39%) and High (>40%) sTILs scores as imaged on OptraScan at 10X magnification and processed using ImageViewer 2.0.4. Blue lines are the scale bars representing 200um. **B**. Bar graph demonstrating percent frequency across binned categories of sTILs scores in ER+, HER2+ and TNBC subtypes. The number of patients in each bin (n) are shown on top of the bars. Association of categorical sTILs variable (Low, Moderate and High) across breast cancer subtypes was tested for significance by 3*3 χ2 test using GraphPad Prism v.5.

**Table S2:** Response to NACT across breast cancer subtypes. Number of patients treated with NACT and their response to NACT is recorded as complete, partial, no response as in stable disease or progressive disease. Number of patients in each response category for ER+, HER2+ and TNBC are given in the table. The distribution across subtypes for pCR status was tested using χ2 test and was found to be non-significant.

|  |  | Total | ER+ | HER2+ | TNBC | *p-values* |
| --- | --- | --- | --- | --- | --- | --- |
| **Pos-NACT**<br>**response**<br>**(n=33)** | Complete | 8 | 1 | 1 | 6 | 0.3552 |
|  | Partial | 16 | 9 | 2 | 5 |  |
|  | Stable disease | 8 | 3 | 2 | 3 |  |
|  | Progressive<br>disease | 1 | 1 | 0 | 0 |  |

